# Screening for dementia: Q^*^ index as a global measure of test accuracy revisited

**DOI:** 10.1101/2020.04.01.20050567

**Authors:** Andrew J Larner

## Abstract

Receiver operating characteristic (ROC) curves intersect the downward diagonal through ROC space at a point, the Q* index, where by definition sensitivity and specificity are equal. Aside from its use in meta-analysis, Q* index has also been suggested as a possible global parameter summarising test accuracy of cognitive screening instruments and as a definition for optimal test cut-off. Area under the ROC curve (AUC ROC) is a recognised measure of test accuracy. This study compared different methods for determining Q* index (both graphical and calculation from diagnostic odds ratio) and AUC ROC (integration and calculation from diagnostic odds ratio) using the dataset of a prospective screening test accuracy study of the Mini-Addenbrooke’s Cognitive Examination. The different methods did not agree. DOR-based calculations gave a very sensitive cut-off but with poorer global metrics than the graphical method. DOR-based calculations are not recommended for defining optimal test cut-off or test accuracy.

## Introduction

Screening for dementia in clinical practice usually involves the administration of at least one cognitive screening instrument. The definition of test cut-off or dichotomisation point for such instruments has profound implications for screening accuracy.

Optimal test cut-off or decision threshold may be defined in various ways based on a receiver operating characteristic (ROC) plot of the “hit rate” or Sensitivity or true positive rate (TPR) (ordinate) against the false positive rate (FPR), or [1 – Specificity], (abscissa) across the range of possible test scores. Of these methods, the Euclidean index and the Youden index are probably the most commonly used to define optimal cut-off.

The Euclidean index (d) is the point minimally distant from the top left hand (“north west”) corner of the ROC plot, with coordinates (0,1), with the ROC intersect of a line joining these points designated c,^1,2^ such that:

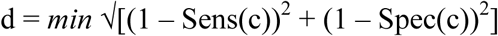

Youden index (Y) is a global or unitary measure of test performance given by [Sensitivity + Specificity – 1]. Maximal Y corresponds to the point on the ROC curve which is the maximal vertical distance from the diagonal line (y = x, or Sens = 1 – Spec, or TPR = FPR), the line representing the performance of a random classifier. Also known as c*, this point minimizes misclassification:^3^

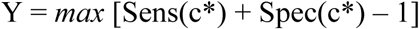

When c and c* do not agree, c* is preferable since it maximizes the overall rate of correct classificiation.^4^

Using a summary ROC (SROC) curve based on data from meta-analyses, another index was proposed to determine optimal test cut-off: the Q* index. This is the point where the downward, negative, or anti-diagonal line through ROC space (y = 1 – x, or Sens = Spec, or Sens – Spec = 0, or TPR = 1 – FPR) intersects the ROC curve.^5^ Q* index may also be defined as the “point of indifference on the ROC curve”, where the sensitivity and specificity are equal, or, in other words, where the probabilities of incorrect test results are equal for disease cases and non-cases (i.e. indifference between false positive and false negative diagnostic errors, with both assumed to be of equal value). For the particular case of a symmetrical ROC curve, Q* will be equal to the Euclidean index.^6^ Q* index has also been suggested as useful global measure of test accuracy,^7^ although its use for the comparison of diagnostic tests is controversial and has been generally discouraged.^8^

As well as being assessed graphically, as the point of intersection of the ROC curve and the anti-diagonal through ROC space, Q* may also be calculated based on its relationship to the diagnostic odds ratio (DOR). DOR, or the cross-product ratio, is the ratio of the product of true positives and true negatives and of false negatives and false positives.^9,10^ Q* may be calculated as:^6^

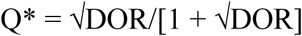

In addition to defining appropriate cut-offs, clinicians are often interested in the overall accuracy of a test. This may also be measured from the ROC curve as the area under the curve (AUC), typically calculated by integration. AUC may also be calculated, based on its relationship to DOR:^6^

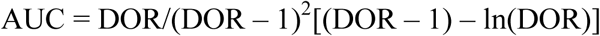

Comparison of Q* index and AUC values determined by these different methods has not, to the author’s knowledge, been previously examined.

Therefore, the aims of the current study were:

- To define and compare optimal cut-offs using the ROC-based indices d, Y, and Q*, the latter assessed both graphically and by calculation using DOR.
- To use the optimal cut-offs thus defined to examine the implications for test sensitivity and specificity and predictive values.
- To compare AUC values for test accuracy calculated both by integration and by calculation using DOR.

These aims were realised by reanalysing the dataset of a large pragmatic prospective screening test accuracy study of the Mini-Addenbrooke’s Cognitive Examination (MACE),^11^ a cognitive screening instrument widely used in the assessment of patients suspected to have dementia and lesser degrees of cognitive impairment. MACE comprises tests of attention, memory (7-item name and address), verbal fluency, clock drawing, and memory recall, takes around 5-10 minutes to administer, and has a score range 0-30 (impaired to normal). In the index MACE study, two cut-off points were identified: ≤25/30 had both high sensitivity and specificity (both ≥ 0.85); and ≤21/30 had high specificity (1.00).^12^ The current study builds on previously reported data from the pragmatic prospective test accuracy study of MACE.^13^

## Methods

Optimal MACE cut-off as defined by minimal Euclidean index (d), by maximal Youden index (Y), and by Q* index was determined. Plots of d and Y against MACE cut-offs were used to determine their optima.^13^ Q* index was measured graphically, by reading off the point of intersection of the ROC plot with the anti-diagonal.^7,11^

Q* index was also calculated at each MACE cut-off, encompassing the range of MACE cut-offs defined in the index study,^12^ from the corresponding value of DOR,^11^ using Walter’s formula:^6^

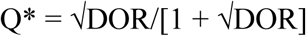

At each of the “optimal” cut-offs thus defined by d, Y, and Q* index, test sensitivity, specificity, and predictive values were then calculated and the results compared. Clinical utility indexes (CUI+, CUI-), parameters indicative of the value of a diagnostic method for ruling in or ruling out a diagnosis, were also calculated and classified,^14^ where:

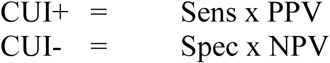

From these, the global summary screening utility index (SSUI) was calculated and classified:^15^

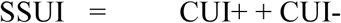

Test accuracy was assessed by means of the area under the ROC curve (AUC). AUC was calculated by the standard method of integration with 95% confidence intervals, and also by calculation, using Walter’s formula:^6^

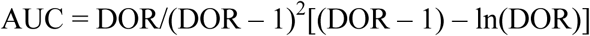

## Results

Optimal MACE cut-off differed according to method. Plots of d and Y versus MACE test cut-offs showed minimum d at ≤19/30, and maximum Y at ≤20/30. Graphically, Q* index was 0.8 at cut-off ≤18/30 (Figure 1).

**Figure 1:**
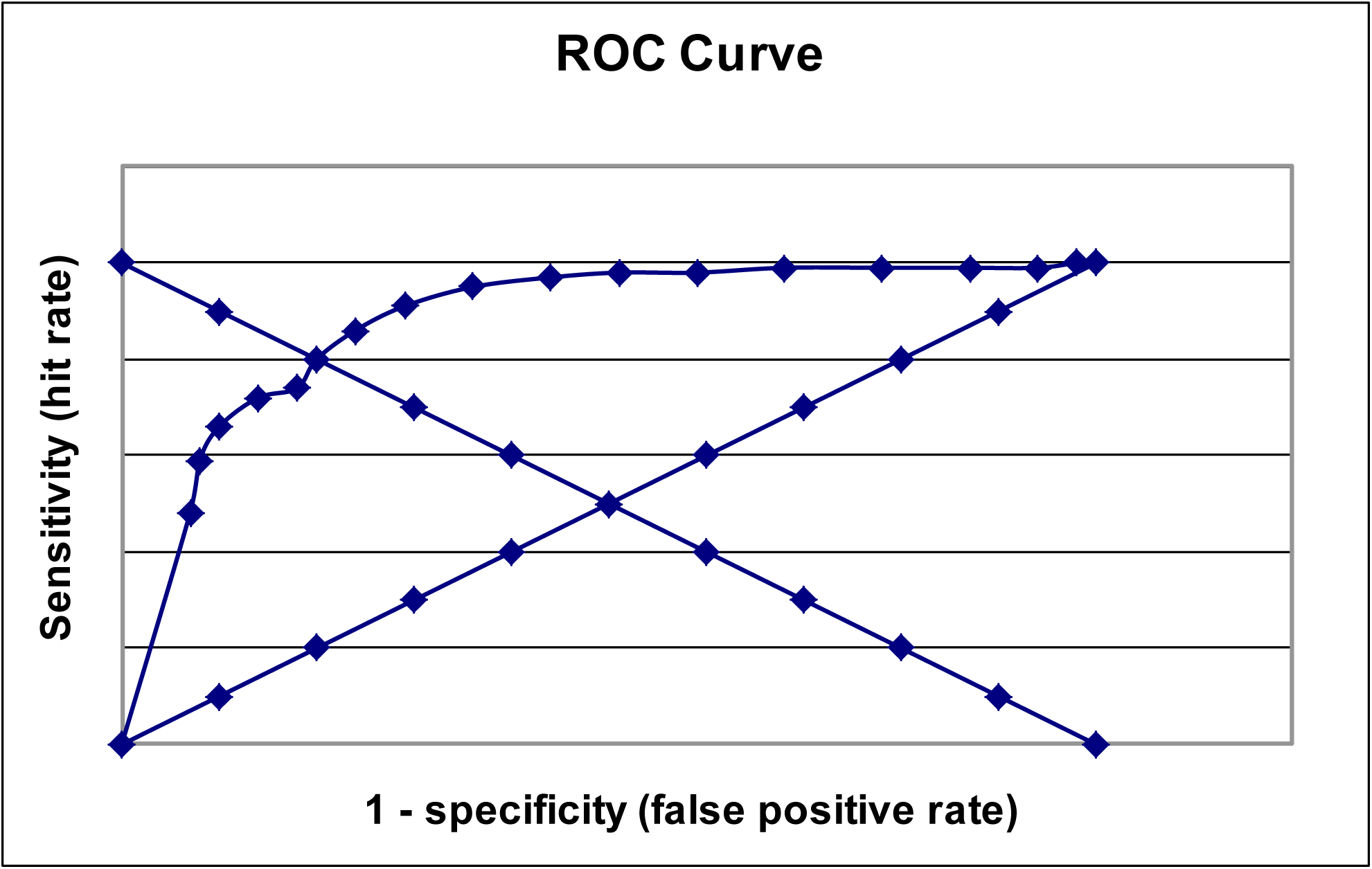
MACE ROC plot for diagnosis of dementia, with chance diagonal (y = x) and anti-diagonal (y = 1 – x) lines, the latter permitting the graphical estimate of Q* index.

By calculation, Q* index varied with diagnostic OR at each test cut-off (Table 1, 3^rd^ column), with the optimal value at ≤23/30. Of note, the calculated Q* index at cut-off ≤18/30 corresponded with the value (0.8) determined graphically.

**Table 1:** Q* index and AUC values calculated from diagnostic odds ratio at each MACE cut-off value

| **MACE cut-off** | **DOR** | **Calculated Q\* index** | **Calculated AUC** |
| --- | --- | --- | --- |
| ≤25/30 | 52.0 | 0.878 | 0.941 |
| ≤24/30 | 38.7 | 0.862 | 0.927 |
| ≤23/30 | 52.8 | 0.879 | 0.941 |
| ≤22/30 | 47.7 | 0.873 | 0.937 |
| ≤21/30 | 32.2 | 0.850 | 0.917 |
| ≤20/30 | 25.1 | 0.834 | 0.902 |
| ≤19/30 | 18.9 | 0.813 | 0.882 |
| ≤18/30 | 16.0 | 0.800 | 0.869 |
| ≤17/30 | 12.8 | 0.782 | 0.850 |

Abbreviations: DOR = diagnostic odds ratio; AUC = area under the ROC curve

Comparison of test sensitivity, specificity, and predictive values at each of the defined “optimal” cut-offs showed variations in all parameters, with the exception of NPV which remained fairly constant (Table 2). Sensitivity and specificity were not equal for the calculated Q* index, unlike the graphically determined Q* index.

**Table 2:**
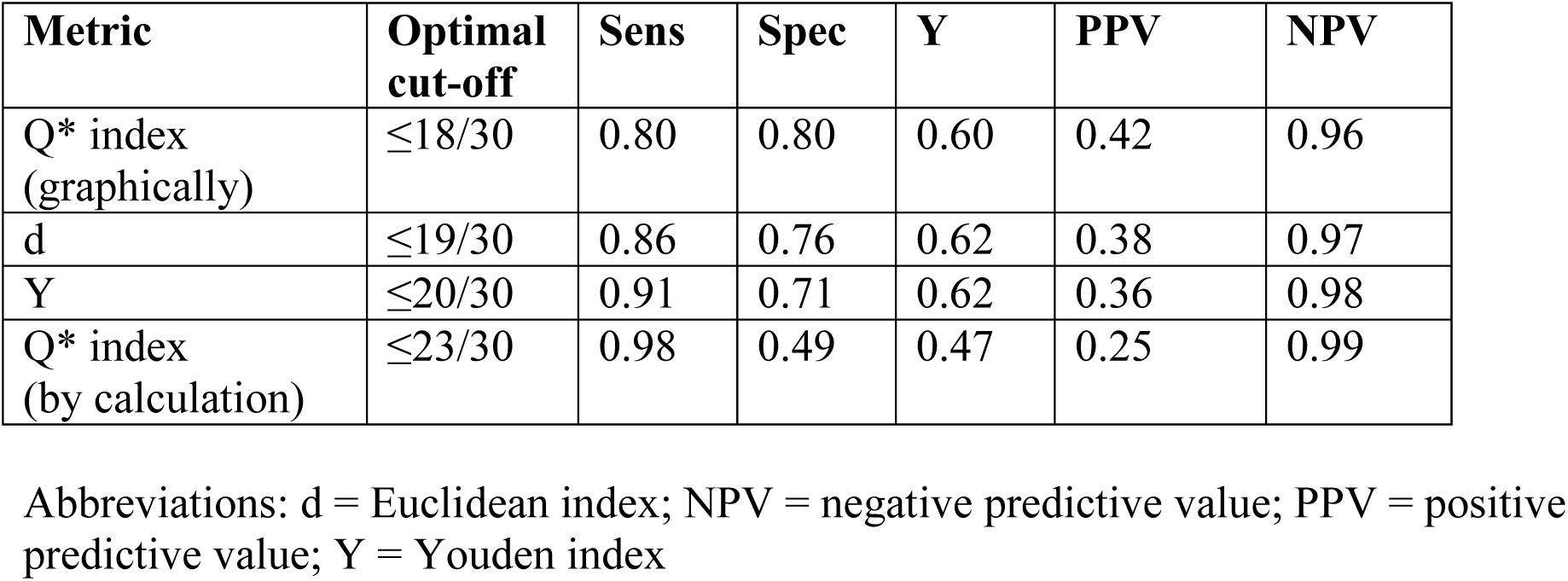
MACE sensitivity, specificity, Youden index, and predictive values at optimal cut-offs defined by various global metrics

**Table 3:** MACE clinical and screening summary utility indexes and their classification at the optimal cut-offs defined by various global metrics

| **Metric** | **Optimal cut-off** | **CUI+** | **CUI-** | **SSUI** |
| --- | --- | --- | --- | --- |
| Q* index (graphically) | $\leq 18/30$ | 0.336<br>(very poor) | 0.768<br>(good) | 1.104<br>(adequate) |
| d | $\leq 19/30$ | 0.327<br>(very poor) | 0.737<br>(good) | 1.064<br>(adequate) |
| Y | $\leq 20/30$ | 0.378<br>(poor) | 0.696<br>(good) | 1.023<br>(adequate) |
| Q* index (by calculation) | $\leq 23/30$ | 0.245<br>(very poor) | 0.485<br>(poor) | 0.730<br>(poor) |

Abbreviations: d = Euclidean index; CUI+, CUI- = positive and negative clinical utility index; SSUI = summary screening utility index; Y = Youden index

Clinical utility indexes and summary screening utility index values for each cut-off method showed the calculated Q* index to have the poorest performance of the methods examined (Table 3).

AUC by integration was 0.886 (95% confidence interval 0.856-0.916).^11^ Calculated AUC showed a maximum of 0.941 at the optimal cut-off of ≤23/30 (Table 1, 4^th^ column). The calculated AUC most closely corresponding to the AUC by integration was between the cut-offs of ≤19/30 and ≤20/30, the optima defined by d and Y respectively.

## Discussion

Optimal cut-off values defined by different ROC-based methods differ, and this has implications for test performance in terms of the sensitivity, specificity, and predictive values calculated from these cut-offs.

Symmetrical ROC curves have constant DOR. Hence for the ROC diagonal line (y = x, or Sens = 1 – Spec, or TPR = FPR), DOR = 1, indicative of a useless test. In an asymmetric ROC plot, the more usual situation in clinical practice (Figure 1), DOR varies with test cut-off or threshold, with corresponding variation in the value of Q* index calculated using Walter’s equation. For the anti-diagonal line (y = 1 – x, or Sens = Spec, or Sens – Spec = 0, or TPR = 1 – FPR), DOR will vary from ∞ at coordinates (0,1) through 1 at the intersection with the ROC diagonal (Sens = Spec = 0.5) to -∞ at the bottom right hand (“south east”) corner at coordinates (1,0).

In the current study, the optimal MACE cut-off defined by Q* index by calculation from DOR appeared to be an outlier compared to the other methods examined for defining optimal cut-off. This confirmed the findings of a previous study looking at the use of various global measures to define MACE cut-offs where optimal cut-off defined by DOR was an outlier.^13^ This higher cut-off gave better sensitivity but poorer specificity and positive predictive value than the other defined “optimal” cut-offs (Table 2). Furthermore, the AUC by calculation from DOR gave a higher value of AUC than by integration.

Clinicians desirous of using highly sensitive tests to avoid missing cases of dementia, even at the cost of more false positives, might therefore be inclined to use the DOR-based method to define test cut-off. However, it is recognised that DOR tends to give the most optimistic results by choosing the best quality of a test and ignoring its weaknesses, producing extremely high cut-points and treating false negatives and false positives as equally undesirable. Moreover, DOR may be less intuitive as a test measure than sensitivity and specificity.^8^ It may be noted that, of the methods examined here, Q* index by calculation from DOR gave the lowest value for the Youden index (Table 2), and for clinical utility indexes and the summary screening utility index (Table 3).

Hence this study provides additional evidence of the shortcomings of DOR and DOR-based calculations^16,17^ and its use cannot therefore be recommended for defining optimal test cut-off or test accuracy.

## Data Availability

Data available from author

## References

[1] Zweig MH, Campbell G. Receiver-operating characteristic (ROC) plots: a fundamental evaluation tool in clinical medicine. Clin Chem 39: 561–577 (1993).

[2] Coffin M, Sukhatme S. Receiver operating characteristic studies and measurement errors. Biometrics 53: 823–837 (1997).

[3] Schisterman EF, Perkins NJ, Liu A, Bondell H. Optimal cut-point and its corresponding Youden Index to discriminate individuals using pooled blood samples. Epidemiology 16: 73–81 (2005).

[4] Zhou XH, Obuchowski NA, McClish DK. Statistical methods in diagnostic medicine (2nd edition). Hoboken, N.J.: John Wiley; p. 51–52 (2011).

[5] Moses LE, Shapiro D, Littenberg B. Combining independent studies of a diagnostic test into a summary ROC curve: data-analytic approaches and some additional considerations. Stat Med 12: 1293–1316 (1993).

[6] Walter SD: Properties of the summary receiver operating characteristic (SROC) curve for diagnostic test data. Stat Med 21: 1237–1256 (2002).

[7] Larner AJ. The Q* index: a useful global measure of dementia screening test accuracy? Dement Geriatr Cogn Dis Extra 5: 265–270, doi: 10.1159/000430784 (2015).

[8] Lee J, Kim KW, Choi SH, Huh J, Park SH. Systematic review and metaanalysis of studies evaluating diagnostic test accuracy: a practical review for clinical researchers – Part II. Statistical methods of meta-analysis. Korean J Radiol 16: 1188–1196, doi: 10.3348/kjr.2015.16.6.1188 (2015).

[9] Edwards AWF. The measure of association in a 2×2 table. J R Stat Soc Ser A 126: 109–114 (1963).

[10] Glas AS, Lijmer JG, Prins MH, Bonsel GJ, Bossuyt PM. The diagnostic odds ratio: a single indicator of test performance. J Clin Epidemiol 56: 1129–1135 (2003).

[11] Larner AJ. MACE for diagnosis of dementia and MCI: examining cut-offs and predictive values. Diagnostics (Basel) 9: E51, doi: 10.3390/diagnostics9020051 (2019).

[12] Hsieh S, McGrory S, Leslie F, Dawson K, Ahmed S, Butler CR, et al. The Mini-Addenbrooke’s Cognitive Examination: a new assessment tool for dementia. Dement Geriatr Cogn Disord 39: 1–11, doi: 10.1159/000366040. (2015).

[13] Larner AJ. Defining “optimal” test cut-off using global test metrics: evidence from a cognitive screening instrument. Neurodegener Dis Manag 10: accepted (2020).

[14] Mitchell AJ. Sensitivity x PPV is a recognized test called the clinical utility index (CUI+). Eur J Epidemiol 26: 251–252, doi: 10.1007/s10654-011-9561-x. (2011).

[15] Larner AJ. New unitary metrics for dementia test accuracy studies. Prog Neurol Psychiatry 23(3): 21–25, doi: 10.1002/pnp.543 (2019).

[16] Bohning D, Holling H, Patilea V. A limitation of the diagnostic-odds ratio in determining an optimal cut-off value for a continuous diagnostic test. Stat Methods Med Res 20: 541–550, doi: 10.1177/0962280210374532 (2011).

[17] Hajian-Tilaki K. The choice of methods in determining the optimal cut-off value for quantitative diagnostic test evaluation. Stat Methods Med Res 27: 2374–2383, doi: 10.1177/0962280216680383 (2018).

